# Dual-Filament 3D Printing of Patient-Specific CT Phantoms with Embedded Implants and Tunable Metal-Artifact Intensity

**DOI:** 10.64898/2026.07.17.26358319

**Authors:** Pouyan Pasyar, Kai Mei, Jessica Y. Im, Leonid Roshkovan, Michael Geagan, Peter B. Noël

**Affiliations:** Department of Radiology, Perelman School of Medicine, University of Pennsylvania, Philadelphia, PA, USA; Department of Bioengineering, School of Engineering and Applied Science, University of Pennsylvania, Philadelphia, PA, USA

**Keywords:** 3D printing methodology, fused deposition modeling, voxel-level multi-material printing, spectral computed tomography, metal artifact reduction, phantom design, orthopedic implants, dual filament, PixelPrint

## Abstract

**Background:** Metallic implants such as orthopedic screws, prostheses, and dental hardware produce beam-hardening, photon-starvation, and streak artifacts that degrade computed tomography (CT) image quality, and the metal artifact reduction (MAR) methods developed to mitigate them require objective, reproducible benchmarking.

**Purpose:** Objective evaluation of MAR algorithms in CT is hindered by the absence of phantoms that simultaneously provide anatomically realistic backgrounds, embedded implants of known geometry, and controllable, ground-truth–referenced artifact intensity. We present a dual-filament, voxel-level three-dimensional (3D) printing method that fulfills these requirements and demonstrate its capabilities on a clinically representative cervical spine case with embedded orthopedic spinal screws.

**Methods:** The proposed method extends the *PixelPrint* framework, a fused-deposition-modeling (FDM) workflow that converts clinical Digital Imaging and Communications in Medicine (DICOM) data directly into 3D-printer Geometric code (G-code) without intermediate segmentation or surface meshing, to interleaved, voxel-level deposition of two filaments: a calcium-doped polylactic acid (PLA) for soft tissue and bone, and a higher-attenuation metal-doped PLA for metallic implants. For demonstration, anonymized DICOM data of a healthy cervical spine were used to design and fabricate three matched phantoms, each with six embedded spinal screws at C4–C6: a 0% metal-infill ground-truth phantom, a 50% medium-metal-infill phantom, and an 85% high-metal-infill phantom. All phantoms were scanned on a clinical spectral CT system at 120 kVp and 1000 mAs, reconstructed at 0.67 mm slice thickness with virtual monoenergetic imaging (VMI) across 50–190 keV. Method performance was characterized by region of interest (ROI)-based Hounsfield Unit (HU) agreement with the source patient data and by the noise-independent Gumbel-distribution p-index metric.

**Results:** The dual-filament method reproduced patient anatomy, soft-tissue contrast, and screw geometry with high fidelity. ROI HU values agreed with patient data within ±25 HU for soft tissue and trabecular bone; cortical regions were underestimated owing to the current ceiling of the calcium-doped PLA used in this study. The tunable-artifact behavior was quantified as follows: the Gumbel location parameter μ scaled monotonically from 46.7 HU (no-metal background) to 57.1 HU (50% infill) to 90.5 HU (85% infill) for the VMI 70 keV with standard filter. High-keV VMI reconstructions substantially reduced streak and beam-hardening artifacts while preserving anatomic detail.

**Conclusions:** The proposed dual-filament, voxel-level *PixelPrint* method enables the fabrication of patient-specific, multi-material CT phantoms with embedded metallic implants and controllable, ground-truth–referenced artifact intensity. Although demonstrated here in a single cervical-spine case, the workflow is anatomy- and implant-agnostic by construction and could in principle be adapted to other musculoskeletal sites (e.g., knee, hip, dental) and implant materials, providing a reproducible methodological foundation for benchmarking MAR algorithms, characterizing spectral CT performance, and validating emerging photon-counting detector systems.

## 1. INTRODUCTION

Computed tomography (CT) is among the most widely used imaging modalities in modern medicine, supporting diagnosis, surgical planning, and treatment monitoring across virtually every body region. Its diagnostic utility, however, is frequently degraded by metallic implants such as orthopedic screws, prosthetic joints, dental hardware, and surgical clips, which cause beam hardening, photon starvation, and streaking artifacts that obscure adjacent anatomy and reduce diagnostic confidence^1–4^. With the prevalence of spinal and joint surgery rising^5^, metal-induced artifacts represent a growing portion of routine CT workload and a recurrent source of non-diagnostic image regions. A broad spectrum of metal artifact reduction (MAR) techniques has been developed, including sinogram-based interpolation, iterative MAR, high-keV virtual monoenergetic imaging (VMI), and, more recently, deep-learning–based reconstruction approaches^6–11^. Yet each of these techniques can introduce secondary trade-offs, including blurring of adjacent structures, the appearance of new pseudo-artifacts in certain anatomies, or inaccurate Hounsfield Unit (HU) values that bias downstream quantitative tasks^2,6,8^.

Rigorous evaluation and inter-comparison of MAR techniques therefore depend critically on the availability of physical phantoms that meet three combined requirements: (i) anatomically realistic, patient-derived backgrounds with tissue-appropriate attenuation and texture; (ii) embedded metallic implants of known, reproducible geometry; and (iii) a controllable, ground-truth–referenced means of modulating artifact severity. No single existing phantom platform meets all three. Commercial calibration phantoms are invaluable for routine quality assurance but use homogeneous backgrounds and simplified geometries that cannot capture the heterogeneous attenuation, fine textures, and spectral behavior of real human tissues, especially near metal. Anthropomorphic phantoms address anatomical realism only partially, as most rely on uniform resins or gels that cannot reproduce voxel-level attenuation distributions or tissue-specific spectral responses. Phantoms developed specifically for MAR evaluation have historically been either highly simplified or limited in anatomical fidelity; for example, Jeong et al. fabricated an agar-based abdominal phantom containing two sets of spinal screws within a tissue-mimicking medium^12^. Findings on such phantoms may not generalize reliably to clinical practice.

Three-dimensional (3D) printing has emerged as a transformative tool in medical imaging research, enabling the fabrication of cost-efficient, customizable, patient-specific phantoms across CT, MRI, PET, SPECT, and ultrasound^13,14^. However, the dominant 3D printing strategies for CT phantoms have important methodological limitations relative to the three requirements above. Approaches built on segmentation and surface meshing of computer-aided design (CAD) models reproduce gross anatomy but discard the voxel-level attenuation distribution and texture of the source image; single-material fused-deposition-modeling (FDM) approaches can emulate one tissue class but cannot simultaneously represent soft tissue, bone, and a higher-Z implant within a single print; and most prior work treats implant attenuation as a fixed property of the chosen filament rather than as a tunable parameter, precluding direct, paired comparisons of MAR performance across matched anatomy. Our group has previously developed *PixelPrint*, an FDM framework that directly converts Digital Imaging and Communications in Medicine (DICOM) images into 3D-printer Geometric code (G-code) without intermediate segmentation or surface meshing, preserving the texture and attenuation of the original image at the voxel level^15–17^. By modulating filament line widths and deposition speed, *PixelPrint* controls local HU values with high fidelity, and has been validated for lung, soft-tissue, and bone phantoms on both single-energy and spectral CT systems, as well as for deep-learning–reconstruction evaluation, respiratory motion applications, and PET imaging^17–20^. These prior implementations, however, used only a single filament per print and therefore could not address the three combined requirements set out above.

Here we present a dual-filament, voxel-level 3D printing method that resolves these limitations by interleaving two filaments, a calcium-doped polylactic acid (PLA) for soft tissue and bone, and a metal-doped PLA for metallic implants, on a per-voxel basis within a single print, while preserving the patient-derived attenuation and texture of the source DICOM image. Per-voxel filament assignment, line width, and deposition speed jointly modulate local HU and metal-induced artifact severity in a controlled, anatomy-invariant manner. By construction, the method is implant- and anatomy-agnostic: any region of interest in a patient CT scan can be augmented with one or more digitally embedded implant models, and the metal-doped filament infill at the implant locations can be tuned to span a controlled, monotonic range of artifact intensities.

We describe the proposed method and demonstrate its capabilities using a clinically representative case study: three matched, patient-specific cervical spine phantoms, each with six embedded orthopedic spinal screws at C4–C6, printed at 0%, 50%, and 85% metal-doped filament infill. We characterize the phantoms with quantitative ROI-based HU comparison against the source patient data, and with the noise-independent Gumbel p-index artifact metric^21^. We further illustrate the method’s utility for evaluating spectral CT-based MAR strategies by imaging the three phantoms at VMI 50–190 keV (50, 70, 90, 110, 130, and 190 keV).

## 2. MATERIALS AND METHODS

### 2.1 Overview of the dual-filament voxel-level *PixelPrint* method

The proposed method extends the single-filament *PixelPrint* workflow^15–17^ to a dual-filament, voxel-level scheme suitable for multi-material phantoms with embedded high-Z structures. Three properties define the method. First, voxel-level translation: the input DICOM volume is converted directly into 3D-printer G-code without segmentation or surface meshing, so that the patient-derived attenuation and texture of the source image are preserved at the voxel scale. Second, per-voxel filament assignment: two separate voxel arrays are generated per phantom, one assigned to a calcium-doped PLA filament (representing soft tissue and bone), and one assigned to a higher-attenuation metal-doped PLA filament (representing metallic implants), and a strictly complementary, interleaved deposition strategy ensures that the two filaments occupy non-overlapping sub-voxel positions within shared regions. Third, joint density modulation: filament line width, deposition speed, and per-voxel infill fraction jointly control local HU, mapping voxel intensities deterministically onto the reconstructed CT attenuation range through partial-volume averaging (filament lines are intentionally smaller than the reconstructed CT voxel).

### 2.2 Demonstration case: cervical spine phantom with embedded spinal screws

To demonstrate the dual-filament method on a clinically representative case, we designed and fabricated three matched, patient-specific cervical spine phantoms with embedded spinal screws. Anonymized clinical CT data of the cervical spine from a healthy adult were used as the anatomical basis for phantom generation, under an institutional review board–approved protocol with a waiver of informed consent for retrospective use of de-identified images. Six spinal screw models (length 15 mm, outer diameter 4 mm; geometry derived from standard cervical screw dimensions) were digitally embedded into vertebral levels C4–C6, with bilateral placement at each level. Screw CAD models were voxelized at the native image resolution and overlaid onto the patient image volume, enabling direct integration of metallic structures with the surrounding osseous and soft-tissue anatomy while preserving clinically realistic trajectories within the vertebral bodies (**Figure 1**). Three matched phantom variants were generated from this combined volume: (i) a ground-truth phantom with no metal filament (0% metal infill at the screw locations), (ii) a medium-metal phantom with screws printed at 50% metal infill, and (iii) a high-metal phantom with screws printed at 85% metal infill.

**Figure 1.**
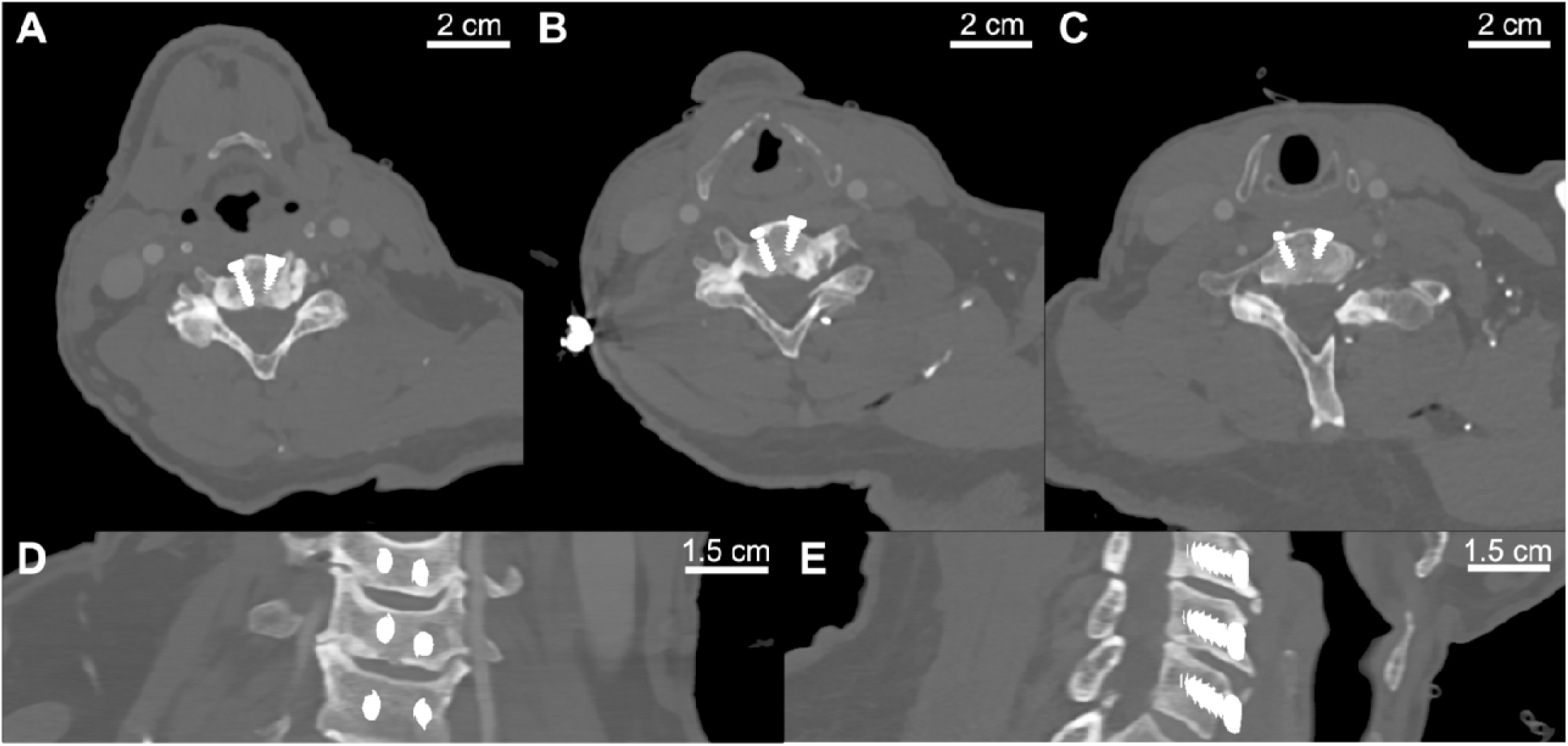
Digital design of the patient-specific cervical spine phantom with six embedded spinal screws at vertebral levels C4 (A), C5 (B), and C6 (C). Axial (A–C), coronal (D), and sagittal (E) views demonstrate anatomically realistic screw placement and alignment. Window level/window width (WL/WW): 0/1500 HU.

The 50% and 85% infill levels were selected as nominal, operationally defined settings spanning a clinically relevant range of artifact severity rather than as calibrated equivalents of specific material. The 50% level produces attenuation and streak characteristics broadly comparable to those of commonly used titanium-based cervical pedicle screws while avoiding the extreme photon starvation of near-solid metallic fills, and the 85% level approximates denser, higher-Z hardware such as cobalt-chromium or stainless steel. This bracketed design allowed systematic, monotonic modulation of artifact severity against an identical anatomical background.

### 2.3 Phantom fabrication

Phantom fabrication used the *PixelPrint* framework, which translates DICOM data directly into 3D-printer G-code without intermediate segmentation or surface meshing, preserving fine image texture and attenuation fidelity by modulating filament line widths at the voxel level. Dual-material phantoms were produced using 1.75 mm calcium-carbonate–doped PLA filament (HDglass / iron-free calcium-doped PLA, Protopasta, Vancouver, WA, USA) to emulate soft tissue and bone, and 1.75 mm metal-doped PLA filament (Ultrafuse 17-4 PH stainless-steel-filled PLA, BASF, Ludwigshafen, Germany) to simulate orthopedic implants. Printing was performed on a dual-extruder FDM 3D-printer (Prusa XL, two-toolhead configuration; Prusa Research, Prague, Czech Republic) equipped with 0.25 mm brass nozzles. Extruder temperatures were 210 °C for the calcium-doped filament and 230 °C for the metal-doped filament, with a bed temperature of 60 °C. Independent extruder calibration in the X–Y plane was verified prior to each print to ensure sub-millimeter co-registration of calcium- and metal-doped filament tracks. Total print time per phantom was approximately 230 hours at 0.2 mm layer height.

### 2.4 Dual-filament 3D printing strategy

The dual-filament printing process used the latest extension of *PixelPrint* to support interleaved multi-material deposition. Two volumetric arrays were generated per phantom: one corresponding to calcium-doped PLA and one corresponding to metal-doped PLA. Each voxel was translated into deposition instructions specifying line width, print speed, and filament assignment, ensuring accurate reproduction of the desired attenuation profile.

Lines were deposited with 90° rotation and lateral shifts between successive layers to minimize directional grid patterns and improve attenuation uniformity. Within each layer containing an implant, calcium- and metal-doped lines were interleaved with a fixed center-to-center line spacing of 0.5 mm, such that one material filled the gaps between adjacent lines of the other in a strictly complementary pattern. Outside the implant region, only calcium-doped filament was deposited; no metal-doped filament was present anywhere in the phantom outside the generated screw volumes.

### 2.5 CT acquisition and image analysis

All phantoms were scanned on a clinical dual-layer spectral CT system (Spectral CT 7500, Philips Healthcare, Cleveland, OH, USA) in helical acquisition mode based on a routine cervical spine protocol. Acquisition parameters included a tube voltage of 120 kVp, a total tube current of 1000 mAs, collimation 64 × 0.625 mm, and rotation time 0.5 s. Images were reconstructed on a 512 × 512 matrix with 0.67 mm slice thickness and 0.34 mm increment using the standard body kernel (B). Spectral base images were reprocessed to generate virtual monoenergetic images (VMIs) at 50, 70, 90, 110, 130, and 190 keV.

Phantom evaluation was performed in three stages: visual quality assessment, quantitative HU analysis, and quantitative artifact analysis. Immediately after fabrication, all phantoms were visually inspected for structural integrity and dimensional accuracy. CT images were then reviewed to confirm faithful reproduction of major cervical-spine landmarks and printed screw trajectories and dimensions were compared against the original CAD models.

For HU analysis, five circular regions of interest (ROIs; nominal area 0.5 cm²) were drawn on a representative axial slice of the cervical spine in the VMI 70 keV reconstruction without MAR for the ground-truth phantom: three in soft-tissue regions (ROIs 1, 2, and 3) and two within osseous structures (ROIs 4 and 5). For each ROI, mean HU and standard deviation were recorded and compared against corresponding ROIs measured on the source patient dataset using the same image-coordinate framework.

To quantify metal artifacts we implemented the robust, noise-independent Gumbel-based p-index, a metric in which streak-artifact severity is summarized by the location parameter (μ) of a Gumbel distribution fitted to the extreme column-wise pixel-intensity differences and normalized against an artifact-free reference, originally described by Cammin et al^21^. Briefly, a rectangular ROI was drawn across a representative streak artifact in the soft tissue dorsal to the C5 spinal screws, and a matched, artifact-free background ROI of identical dimensions was extracted from the same anatomical location in the VMI 70 keV reconstruction for the 0% metal ground-truth phantom. Within each ROI, the largest absolute column-wise pixel-intensity difference was extracted on a per-column basis, the resulting distribution of extreme differences was linearized using a probability-plot transform, and the location parameter μ of a Gumbel distribution was estimated by linear regression. Artifact severity was reported as the location parameter μ for each phantom and as a normalized p-index obtained by comparing the artifact ROI to the matched background ROI. This metric has been shown to be noise-independent and robust to small variations in ROI placement (reported coefficient of variation < 6%), allowing direct comparison across phantoms with different metal content.

Descriptive statistics (mean ± standard deviation) are reported for all quantitative measurements. Phantom-to-patient HU agreement is reported as the absolute mean difference for each ROI. Given the small number of conditions per phantom and the descriptive nature of this proof-of-concept characterization, no inferential statistical tests were performed. Analyses were performed in Python 3.12.7.

## 3. RESULTS

### 3.1 Medium-infill phantom at VMI 70 keV

The 50% metal-infill phantom imaged at VMI 70 keV without MAR (**Figure 2**) closely reproduced the anatomy of the C4–C6 cervical spine. Axial, sagittal, and coronal views show vertebral body morphology, intervertebral spaces, and surrounding soft tissues consistent with the source patient data. The six bilateral spinal screws are clearly visible, with geometry and trajectories matching their digital CAD models. Realistic image textures and soft-tissue contrast are preserved throughout: cortical and trabecular bone are clearly differentiated, the airway is well delineated, and muscle, fat, and adjacent soft-tissue planes are distinguishable across axial slices, with HU values within clinically relevant ranges (see 3.4).

**Figure 2.**
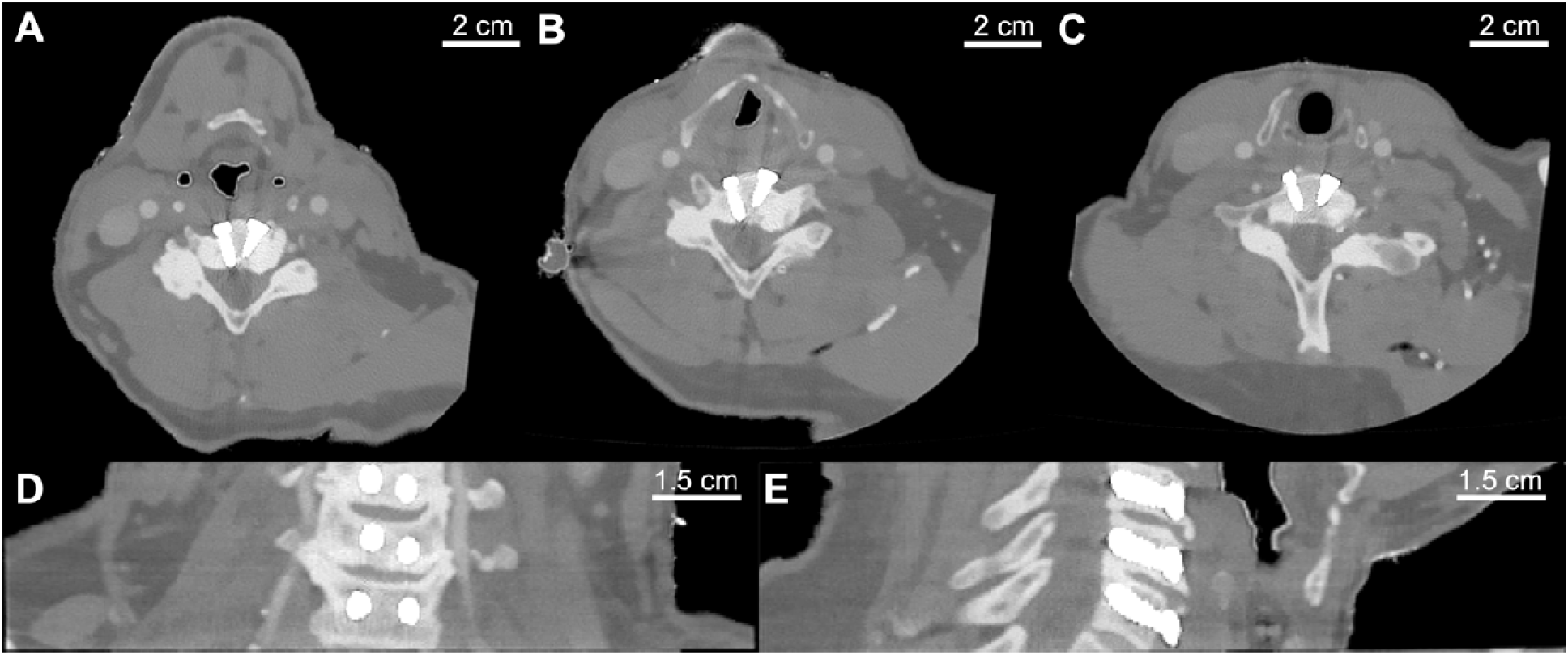
*PixelPrint* cervical spine phantom (50% metal infill) imaged at VMI 70 keV without MAR at vertebral level C4 (A), C5 (B), and C6 (C). Axial (A–C), coronal (D), and sagittal (E) views demonstrate accurate vertebral anatomy, screw geometry, and preserved soft-tissue contrast. WL/WW: 0/1500 HU.

### 3.2 Comparison of 0%, 50%, and 85% metal-infill phantoms at VMI 70 keV

**Figure 3** shows axial, sagittal, and coronal reconstructions of the three phantoms at VMI 70 keV without MAR, demonstrating the effect of metal-doped filament infill on artifact generation. The ground-truth phantom (0% metal) is free of visible streaking or beam-hardening, providing a clean anatomical reference. In the medium-infill phantom (50% metal), mild streak artifacts emerge around the screw interfaces while overall anatomical detail and soft-tissue contrast remain well preserved. In the high-infill phantom (85% metal), pronounced streaks and photon-starvation artifacts radiate from the C4–C6 levels and partially obscure adjacent soft tissues, although the underlying anatomy remains identical to the other two phantoms. This monotonic visual progression is consistent with the intended use of metal infill as a tunable, anatomy-invariant artifact control.

**Figure 3.**
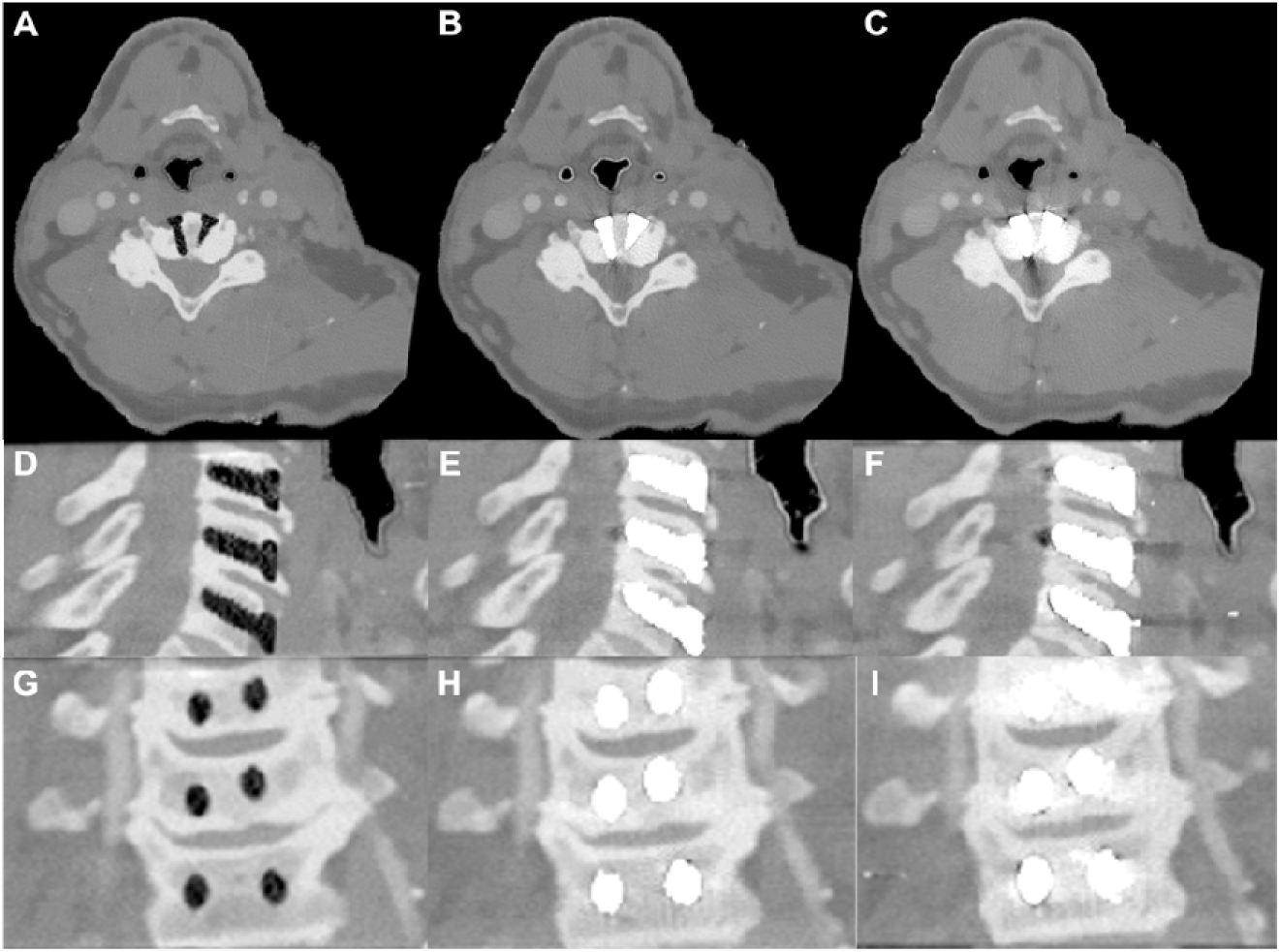
Comparison of three *PixelPrint* cervical spine phantoms imaged at VMI 70 keV without MAR. From left to right, 0%, 50%, and 85% metal infill phantoms are shown in axial (A–C), sagittal (D–F), and coronal (G–I) views. Increasing metal infill results in progressively stronger streak and photon-starvation artifacts while anatomy remains unchanged. WL/WW: 0/1500 HU.

### 3.3 Effect of virtual monoenergetic energy on metal artifacts

To characterize the energy dependence of metal-artifact severity, the three phantoms (0%, 50%, and 85% metal infill) were reconstructed across a virtual monoenergetic image (VMI) sweep at 50, 70, 90, 110, 130, and 190 keV without MAR (**Figure 4**). Two consistent visual trends emerged. Along the infill axis, artifact severity increased from the 0% ground-truth phantom, in which the screw cavities appear as low-attenuation voids with the surrounding anatomy free of streak or beam-hardening artifact at every energy, to the 50% and 85% phantoms, in which the metal-doped implants appear strongly hyperattenuating and generate streak, beam-hardening, and photon-starvation artifacts that are most pronounced at 85% infill. Along the energy axis, increasing the monoenergetic level progressively suppressed these artifacts: at 50 keV the implants showed marked blooming and a bright peri-implant halo together with dark photon-starvation bands (most severe in the 85% phantom), whereas at higher energies (≥90 keV) streak intensity and blooming were visibly reduced, and at 130–190 keV the screw contours were sharply delineated with only faint residual streaking between the screws in the high-infill phantom. Consistent with the known energy dependence of beam hardening and photon starvation, the lowest energy (50 keV) maximized soft-tissue and bone contrast but also maximized metal-artifact severity, whereas high-keV reconstructions traded a flatter overall contrast for substantially cleaner peri-implant regions and improved implant delineation. Because these trends progressed monotonically and predictably along both the infill and energy axes against a fixed anatomical background and a 0% ground-truth reference, the phantom series provides a controlled testbed for evaluating energy-based (VMI) artifact-reduction strategies, in agreement with prior clinical and phantom reports on high-keV VMI for metal-artifact reduction^9^.

**Figure 4.**
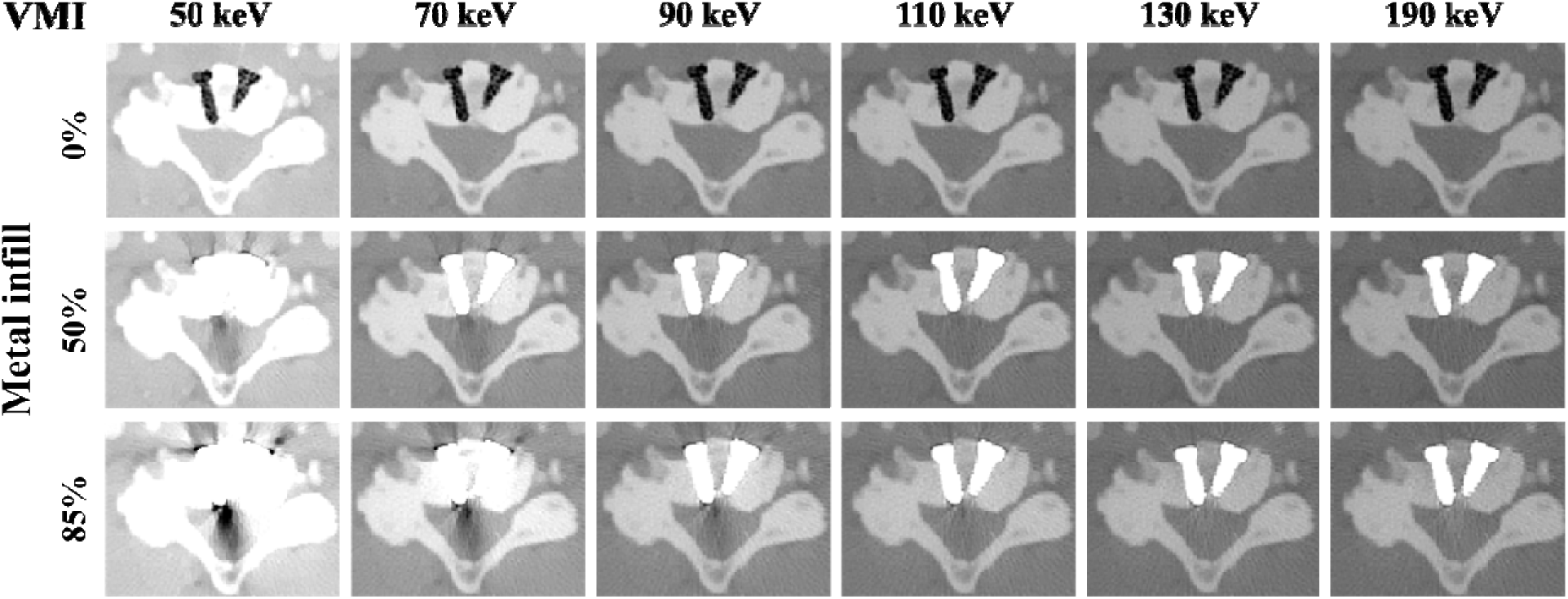
Virtual monoenergetic energy dependence of metal artifacts in the *PixelPrint* cervical spine phantoms. The three phantoms (0%, 50%, and 85% metal infill; rows) are shown at VMI 50, 70, 90, 110, 130, and 190 keV (columns), reconstructed without MAR. Increasing photon energy (left to right) progressively reduces streak, beam-hardening, and photon-starvation artifacts and implant blooming, while increasing metal infill (top to bottom) increases artifact severity. In the 0% ground-truth phantom the screw cavities appear as low-attenuation voids without artifact at all energies. WL/WW: 0/1500 HU.

### 3.4 Quantitative attenuation analysis

Five ROIs (three soft-tissue, two osseous) were placed on a representative axial slice and compared between the phantom and the source patient data (**Figure 5**). In the soft-tissue and trabecular bone regions, mean HU values agreed between the phantom and patient images within ±25 HU (ROIs 1, 2, 4, and 5), confirming that calcium-doped PLA accurately reproduces the attenuation and contrast relationships of soft tissue and moderate-density bone. The largest disagreement occurred in the densest cortical bone region (ROI 4): the patient scan exceeded 1000 HU in this region, while the calcium-doped PLA reached a maximum of approximately 620 HU, reflecting the upper attenuation limit of the current filament formulation. Despite this under-representation in extreme-density areas, the relative bone-to-soft-tissue contrast and tissue texture were well preserved, yielding anatomically realistic image appearance and quantitative fidelity across most structures relevant to MAR evaluation.

**Figure 5.**
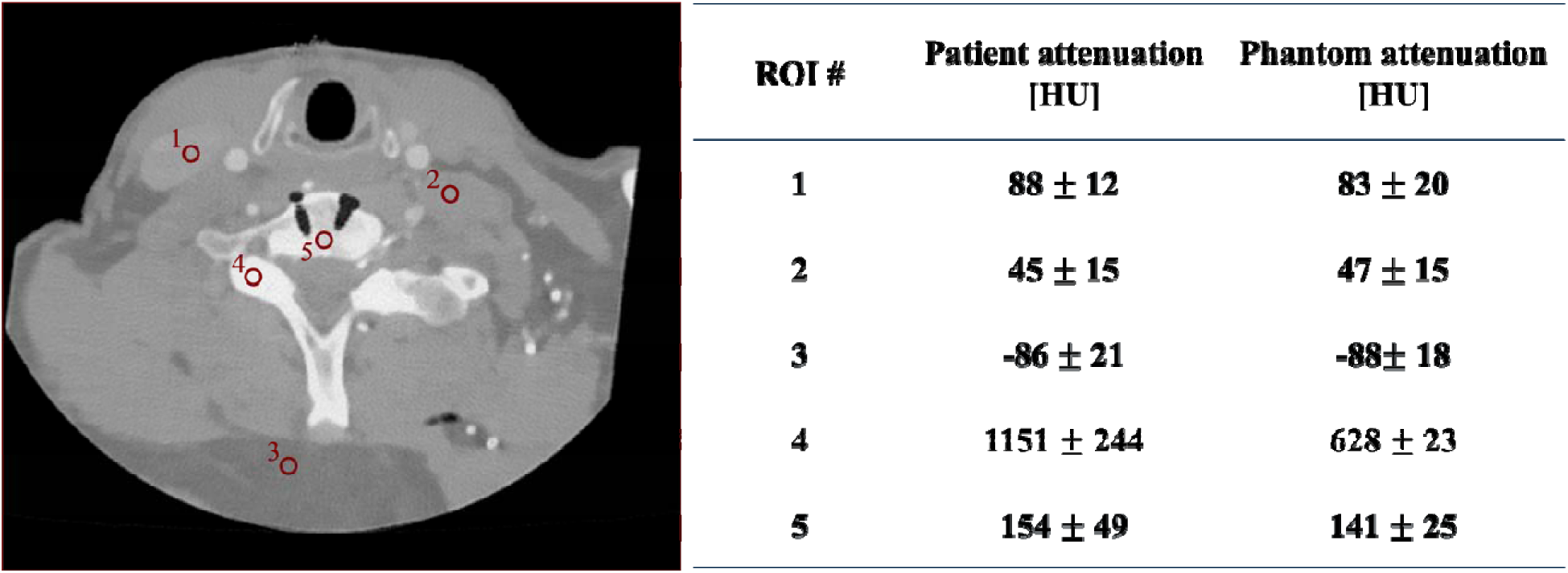
ROI-based evaluation of Hounsfield Unit (HU) accuracy in the *PixelPrint* cervical spine phantom. Representative axial image shows ROI placement, and the accompanying table summarizes mean HU and standard deviation in the phantom relative to the reference patient data. WL/WW: 0/1500 HU.

### 3.5 Quantitative analysis of metal artifacts

Quantitative analysis of the rectangular ROIs drawn beneath the spinal screws showed a monotonic relationship between metal infill and artifact severity (**Figure 6**). The artifact-free background ROI in the ground-truth (0% metal) phantom showed a uniform intensity profile, with a baseline Gumbel location parameter μ = 46.7 HU, consistent with image noise alone. The reported μ values were obtained from a single artifact ROI and one matched background ROI per phantom. In the 50% and 85% metal-infill phantoms, the location parameter increased to μ = 57.1 HU and μ = 90.5 HU, respectively, corresponding to progressively broader distributions of extreme pixel-intensity differences within the soft tissue adjacent to the metal. The linearized Gumbel plots showed decreasing slope with increasing metal content, again consistent with stronger streak intensity gradients around the screws.

**Figure 6.**
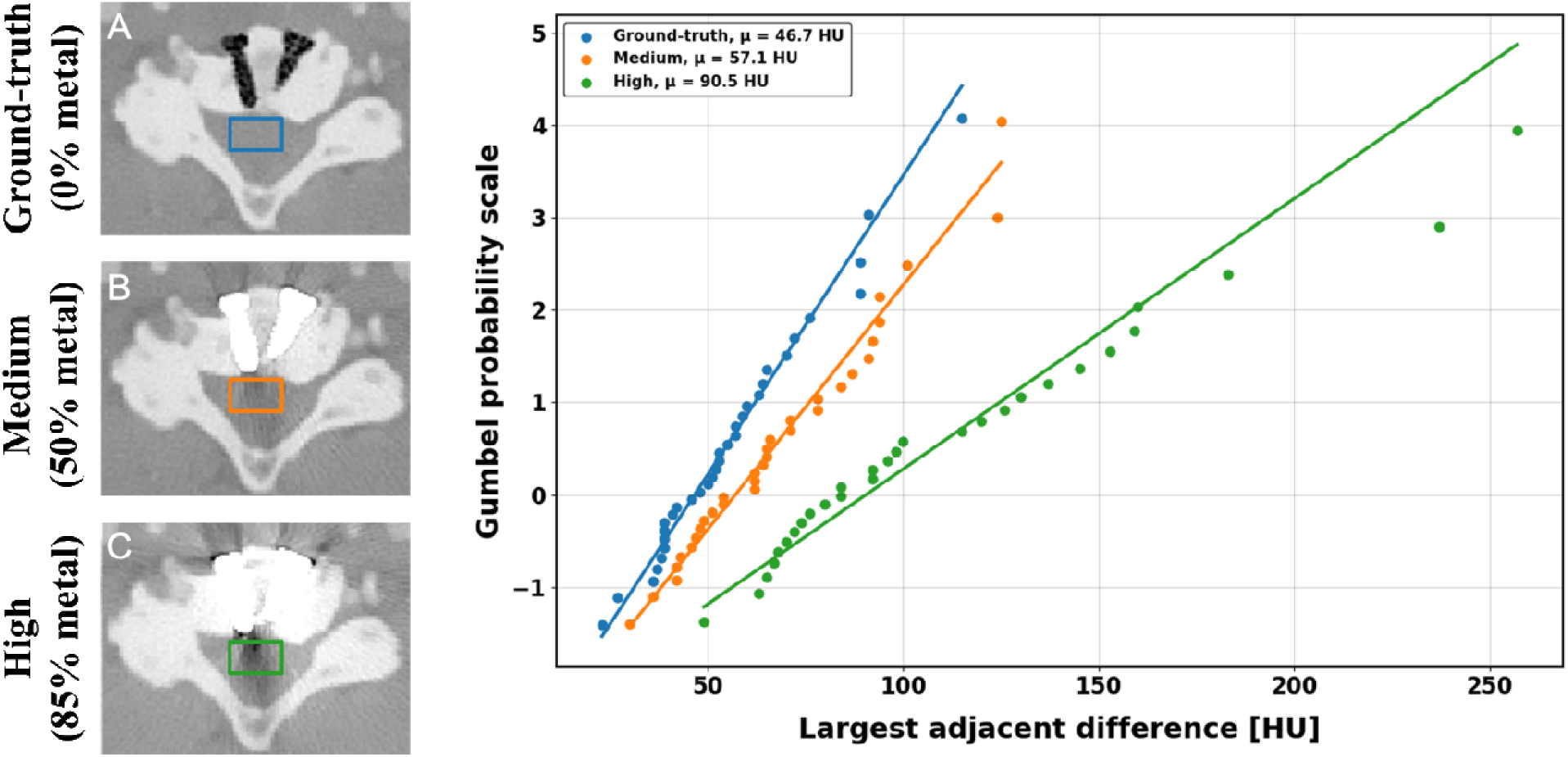
Quantitative analysis of metal-artifact severity using the Gumbel p-index. Axial images show artifact evaluation ROIs for (A) 0%, (B) 50%, and (C) 85% metal-infill phantoms, with corresponding Gumbel distribution fits. The Gumbel location parameter μ increases with metal infill, indicating stronger metal-induced artifacts. WL/WW: 0/1500 HU.

Using the no-metal background ROI as the reference, the normalized p-index increased from 0.252 for the 50% metal phantom to 0.932 for the 85% metal phantom—an approximately 3.7-fold increase in artifact severity over the studied infill range. This indicates that higher metal infill produces stronger streak artifacts in a controlled manner; reproducibility across repeated prints was not assessed, as a single phantom replicate per condition was analyzed. Importantly, because the p-index is noise-independent and robust to small ROI shifts, the observed progression reflects genuine differences in artifact magnitude rather than scan-to-scan noise variability, supporting the use of the Gumbel-based metric for MAR evaluation in *PixelPrint* phantoms.

## 4. DISCUSSION

We have presented a dual-filament, voxel-level 3D printing method for fabricating patient-specific CT phantoms with embedded metallic implants and controllable, ground-truth–referenced artifact intensity. The method extends the *PixelPrint* framework with per-voxel filament assignment and complementary interleaved deposition of a calcium-doped and a metal-doped PLA, while preserving the patient-derived attenuation and texture of the input CT volume. The cervical spine demonstration case validated the three defining capabilities of the method on a clinically representative anatomy. First, the printed phantoms faithfully replicated vertebral morphology, soft-tissue contrast, and screw geometry from the source patient data and digital design. Second, the artifact severity surrounding the implants scaled monotonically with metal-doped filament infill on a clinical spectral CT system, indicating that photon starvation and beam-hardening can be modulated through filament density while the anatomical background is held fixed. Third, holding the anatomy fixed across three matched phantoms enabled paired comparison of MAR-relevant reconstructions against a ground-truth reference, complementing existing MAR phantom designs that rely on simplified geometries or single-condition implants.

Quantitative analysis supported the visual findings. Mean HU values in soft-tissue and cortical bone regions closely matched those in the reference patient scan, generally within ±25 HU; because phantom and patient were imaged in separate acquisitions, ROIs were placed at approximately corresponding locations, leaving some residual registration uncertainty in this comparison. The only notable deviation occurred in the densest trabecular bone, where calcium-doped PLA reached a maximum attenuation of approximately 628 HU, compared with more than 1000 HU in the clinical scan. This ceiling reflects the limitations of the printing material available when the phantoms were fabricated. Further development is underway to better replicate dense bone regions. Despite this ceiling, relative bone-to-soft-tissue contrast was preserved, producing visually and quantitatively realistic images. The Gumbel-based p-index provided an objective, noise-independent measure of artifact intensity that increased consistently with higher metal content, and the normalized p-index spanned 0.25–0.93 across the studied infill range; comparable in dynamic range to clinical streak-artifact severity reported with the same metric in prior work^21^. Together, these findings indicate that *PixelPrint* metal-infill modulation governs artifact magnitude in a predictable manner, offering a reproducible platform for evaluating MAR algorithms under known ground-truth conditions.

Across the virtual monoenergetic sweep (50–190 keV), metal-artifact severity decreased monotonically with increasing photon energy, and the highest energies (130–190 keV) substantially reduced streak artifacts and improved delineation of screw contours and bone–metal interfaces, in agreement with prior reports on the use of high-keV VMI for MAR^7,10^. Our results extend these observations by demonstrating, in matched ground-truth-referenced phantoms, that the magnitude of high-keV artifact reduction tracks with the underlying metal density. This kind of controlled paired comparison is difficult to achieve with patients, where neither the implant density nor the surrounding anatomy can be matched across cases, but is straightforward with *PixelPrint* phantoms.

The method advances prior 3D-printing approaches to CT phantom fabrication along three axes. Compared with single-material *PixelPrint* work, which has primarily focused on lung, soft-tissue, and bone phantoms, the present method adds a second, higher-attenuation material in a strictly complementary, voxel-level interleaved pattern, so that anatomy and implant attenuation can be controlled independently within a single print. Compared with CAD-/mesh-based 3D printing of anthropomorphic phantoms, which preserve gross geometry but discard the voxel-level attenuation distribution of the source patient image, the proposed workflow is segmentation-free and preserves patient-derived texture and HU. Compared with conventional commercial phantoms^22–24^ or with prior MAR-specific phantoms based on water-equivalent gels, epoxy, or isolated metal rods^12,25^, the dual-filament method adds controllable metal density, full patient anatomy, and a ground-truth reference within a single matched series, enabling more direct translation of experimental findings to clinical scenarios.

A central design choice of the method is its anatomy- and implant-agnostic construction. Because each voxel of the input DICOM volume is independently assigned to either the soft-tissue/bone filament or the metal-doped filament, the workflow is not specific to the cervical spine demonstrated here: any anatomical region from a patient CT scan can in principle be augmented with one or more digitally embedded implant models, with the metal-doped filament infill at the implant locations tuned to span the desired range of artifact intensities. Plausible near-term applications include total joint arthroplasty (knee, hip, shoulder), spinal fusion hardware at thoracolumbar levels, dental implants and orthodontic hardware, and surgical clips in the abdomen and pelvis. The approach can also be combined with alternative filaments, e.g., titanium- or cobalt-chromium–equivalent filaments, or higher-density calcium- or hydroxyapatite-doped filaments to extend the bone HU range, without changing the underlying voxel-level deposition strategy.

Several limitations of the method, and of the present demonstration, should be acknowledged. Method limitations: First, the calcium-doped PLA used here cannot yet reproduce the highest cortical-bone HU values observed clinically (∼1000–1500 HU), which may limit absolute fidelity in regions of very dense cortical bone. Second, the spectral behavior of 17-4 PH stainless-steel-doped PLA differs from that of clinical titanium and cobalt-chromium alloys; while the resulting artifact patterns are visually and quantitatively realistic, future work is needed to formally cross-calibrate filament composition against specific implant alloys. Third, in its present implementation the method produces static, non-deformable phantoms, precluding evaluation of motion-related artifacts.

Future work will focus on developing higher-density calcium- or hydroxyapatite-doped filaments to extend the achievable HU range into dense cortical and trabecular bone, incorporating titanium- and cobalt-chromium–equivalent filaments to better match clinical implant materials, and implementing multi-filament *PixelPrint* variants for simultaneous bone, soft-tissue, and contrast-agent simulation.

## 5. CONCLUSION

We have presented a dual-filament, voxel-level 3D printing method that extends the *PixelPrint* framework to multi-material, patient-specific CT phantoms with embedded metallic implants and controllable artifact intensity. By interleaving calcium- and metal-doped PLA filaments at sub-voxel scale under direct DICOM-to-G-code control, the method preserves patient-derived anatomy, soft-tissue contrast, and texture while enabling independent, monotonic modulation of metal-induced artifact severity. Because the underlying workflow is anatomy- and implant-agnostic, the method provides a flexible, reproducible foundation for generating multi-material, implant-containing CT phantoms across musculoskeletal sites and implant chemistries, suitable for the quantitative evaluation of MAR algorithms, spectral CT performance, and emerging photon-counting CT systems^26,27^.

## ACKNOWLEDGEMENTS

We acknowledge support through the National Institutes of Health (R01EB035092, R01EB035908).

## DATA AVAILABILITY STATEMENT

The data that support the findings of this study are available from the corresponding author upon reasonable request.

